# Lateral Thinking: Pathway Specific Neurodegeneration of the Cortical Cholinergic System in Alzheimer’s Disease

**DOI:** 10.1101/2024.07.16.24310492

**Authors:** Rachel A. Crockett, Charlotte Casselton, Tatianna M. Howard, Kevin B. Wilkins, Gang Seo, Helen M. Brontë-Stewart, the Alzheimer’s Disease Neuroimaging Initiative (ADNI) database

## Abstract

**INTRODUCTION:** Atrophy of the nucleus basalis of Meynert (NBM) is an early indicator of Alzheimer’s disease (AD). However, reduced integrity of the NBM white matter tracts may be more relevant for cognitive impairment and progression to dementia than NBM volume. Research is needed to compare differences in NBM volume and integrity of the lateral and medial NBM tracts across early and later stages of AD progression.

**METHODS:** 187 participants were included in this study who were either healthy controls (HC; n=50) or had early mild cognitive impairment (EMCI; n=50), late MCI (LMCI; n=37), or AD (n=50). NBM volume was calculated using voxel-based morphometry and mean diffusivity (MD) of the lateral and medial NBM tracts were extracted using probabilistic tractography. Between group differences in NBM volume and tract MD were compared using linear mixed models controlling for age, sex, and either total intracranial volume or MD of a control mask, respectively. Associations between NBM volume and tract MD with executive function, memory, language, and visuospatial function were also analysed.

**RESULTS:** NBM volume was smallest in AD followed by LMCI (p<0.0001), with no difference between EMCI and HC. AD had highest MD for both tracts compared to all other groups (p<0.001). Both MCI groups had higher lateral tract MD compared to HC (p<0.05). Medial tract MD was higher in LMCI (p=0.008), but not EMCI (p=0.09) compared to HC. Higher lateral tract MD was associated with executive function (p=0.001) and language (p=0.02).

**DISCUSSION:** Integrity of the lateral NBM tract is most sensitive to the earliest stages of AD and should be considered an important therapeutic target for early detection and intervention.

## 1. Introduction

Alzheimer’s disease (AD) is the most common cause of dementia, resulting in 6-7million new cases every year (1). Despite the astronomic global funding for dementia research (US federal government alone invests approximately $3bn annually (2)), there remain no effective treatments available. Consequently, intervening at earlier stages of disease progression remains vital for preserving cognitive function and slowing decline (3, 4). However, greater investigation is needed to determine the brain targets with highest potential for therapeutic benefit.

Mild cognitive impairment (MCI) is the prodromal phase of dementia and is characterised by cognitive impairment beyond what is considered normal for one’s age and education level that does not impact daily living (5). Approximately 5-15% of people with MCI will progress to AD within 10 years of diagnosis (6). Thus, identification of the underlying biomarkers associated with MCI is crucial for early intervention. Significant atrophy of the hippocampus is a pathological criterion for AD (7) and early signs of hippocampal atrophy are also evident in people with MCI (8). However, atrophy of the nucleus basalis of Meynert (NBM) is also a prominent feature (9), which has recently been shown to precede the onset of hippocampal atrophy (10). Therefore, highlighting the potential of this region as an even earlier indicator of disease progression.

Integrity of the NBM white matter pathways have been shown to have more implications for cognitive function than atrophy of the NBM itself (11, 12). There are two main white matter bundles emanating from the NBM. A lateral tract, that supplies the frontal, insula, parietal, and temporal cortices; and a medial tract that supplies the parolfactory, cingulate, pericingulate, and retrospinal cortices (13). Integrity of the lateral tract was significantly reduced in patients with MCI or AD compared to healthy controls (14). Patients with AD also had reduced integrity in the medial tract compared to their healthy counterparts. Tract integrity as measured by mean diffusivity (MD), was most sensitive to disease stage compared to fractional anisotropy or NBM volume. The sensitivity of MD to disease stage is thought to reflect the more accurate nature of neurodegeneration (15, 16), whereas the interpretation of fractional anisotropy may be distorted by the reshaping of white matter tracts being proportional across all diffusion directions (16).

The current definition of MCI encompasses a spectrum of people from those who may be only marginally different to their healthy counterparts to those that are on the cusp of a dementia diagnosis (17). Investigation of patterns of neurodegeneration in these tracts at different stages of MCI is necessary to determine optimal targets for early intervention with the greatest potential to prevent or delay AD onset. In addition, while there is evidence that the lateral tract is associated with measures of executive function, memory and visuospatial function (14), there is limited understanding of the role, if any, of the medial tract in cognition (12, 14, 18). Further elucidating the importance of these tracts for specific cognitive domains is necessary to guide the evaluation of therapeutic targets.

The aim of this study was to investigate differences in NBM volume and white matter integrity of the lateral and medial NBM tracts in healthy controls (HC), people with early MCI, late MCI, and AD. In this context, early versus late MCI is based on severity of cognitive impairment as opposed to timing of disease progression. The secondary aim was to evaluate whether smaller NBM volume and/or poorer integrity of these tracts was associated with poorer cognitive function. We hypothesised that NBM volume and levels of MD of both the lateral and medial NBM tracts would follow disease stage with the highest MD values (worst integrity) evident in AD, followed by the LMCI, EMCI and finally, the HC. In addition, we hypothesised that greater MD of the lateral tract would be associated with poorer executive function, memory, and visuospatial function, while NBM volume and medial tract MD would not be associated with cognitive function.

## 2. Methods

Data used in the preparation of this article were obtained from the Alzheimer’s Disease Neuroimaging Initiative (ADNI) database (https://adni.loni.usc.edu). For up-to-date information on ADNI visit https://adni.loni.usc.edu/.

### 2.1. Participants

A total of 187 participants (50 HC, 50 EMCI, 37 LMCI, and 50 AD) from the ADNI2 cohort were included in this study. The ADNI2 cohort was used due to the inclusion of people with LMCI in this cohort, and to maintain consistent scanning parameters across all scans. There were significantly more participants available in the HC and early EMCI groups. However, to try to maintain similar sample sizes across groups, a subset was randomly selected to match the sample size of the AD group (n=50) for inclusion in the final analyses.

Participants were included in the ADNI data collection if they: 1) were aged 55-90 years old; 2) had a study partner who was willing to provide an independent evaluation of the participant’s functioning; 3) were able to communicate in either English or Spanish; 4) had a geriatric depression score <6; 5) had completed ≥ 6 grades of education; 6) had adequate visual and auditory acuity to undergo neuropsychological testing; 7) were not pregnant, lactating, or of childbearing potential; and 8) had undergone magnetic resonance imaging with both a structural T1-weighted and diffusion weighted scan. They were excluded if they: 1) had any significant neurological disease other than Alzheimer’s disease; 2) had contraindications for MRI; 3) had major depressive disorder, bipolar or a history of schizophrenia; 4) were currently taking psychoactive medications; 5) had a history of alcohol or substance abuse with dependency in the last two years; or 6) had any significant medical illness or laboratory abnormalities that may interfere with the study.

#### 2.1.1. Healthy Controls

Participants were considered a HC if they: 1) scored between 24-30 on the mini-mental state examination (MMSE); 2) scored 0 on the clinical dementia rating (CDR); 3) fell within the education appropriate cut-offs on the logical memory II subscale of delayed recall; and 4) showed no evidence of significant impairment in cognitive functions or activities of daily living.

#### 2.1.2. Mild Cognitive Impairment

Participants were considered to have MCI if they: 1) scored between 24-30 on the MMSE; 2) scored 0.5 on the CDR; 3) scored below the education appropriate cut-offs on the logical memory II subscale of delayed recall; 4) had memory complaints by either the subject or their study partner that is verified by the partner; and 5) showed preserved activities of daily living such that a diagnosis of dementia could not be determined.

They were determined to be either early or late-stage MCI based on education adjusted cut off scores from the Wechsler Memory Scale Logical Memory II. Out of a total score of 25, EMCI was defined as scoring between 9-11 for ≥ 16 years of education; 5-9 for 8-15 years; and 3-6 for ≤ 7 years, while LMCI was defined as scoring ≤ 8 for ≥ 16 years of education; ≤ 4 for 8-15 years; and ≤ 2 for ≤ 7 years.

#### 2.1.3. Alzheimer’s Disease Dementia

Participants were considered to have AD if they: 1) scored between 20-26 on the MMSE; 2) scored 0.5-1 on the CDR; 3) scored below the education appropriate cut-offs on the logical memory II subscale of delayed recall; 4) had memory complaints by either the subject or their study partner that is verified by the partner; and 5) met the NINDS-ADRDA criteria for probable AD.

### 2.2. Cognitive Assessments

The composite z-scores of executive function, memory, language, and visuospatial function were extracted from the ADNI database (19-21). Scores were created using the relevant components of the Alzheimer’s disease assessment scale - Cognitive (ADAS-Cog), Montreal cognitive assessment (MoCA), and MMSE batteries. In addition, executive function also included the WAIS-R digit symbol, digit span forwards and backwards, trail making test parts A and B, and clock drawing test; Memory included the Rey auditory verbal learning test and logical memory; Language included the category fluency, and Boston naming test; and Visuospatial function included the clock copy test.

### 2.3. MRI Acquisition

The MRI scans from the ADNI2 databases were acquired using standardized protocols across sites.

The sequences were acquired on GE Medical Systems Discovery MR750, and Signa HDxt 3T scanners. The T1-weighted (T1-w) images consisted of 200 slices with a 1.2mm^3^ voxel size, an acquisition matrix of 256 x 256, a repetition time (TR) of ∼7000ms, and an echo time (TE) of ∼2.8ms. The diffusion weighted image consisted of 59 slices, with a 2.7mm slice thickness, TR of ∼1300ms, TE of ∼ 70ms, flip angle of 90°, and acquisition matrix of 128 × 128. The *b* = 1000 s/mm^2^ with five *b*0 images, and 41 gradient directions. The images corrected for inhomogeneity, motion, and eddy current corrections were used (22).

#### 2.3.1. Voxel Based Morphometry of NBM

Voxel-based morphometry was applied to T1 images using SPM12 software (Statistical Parametric Mapping 12; Wellcome Trust Centre for Neuroimaging, UCL, London, UK). The T1 images were manually re-orientated to the anterior commissure – posterior commissure line. Images were then automatically segmented into gray matter, white matter, and cerebrospinal fluid with 2mm^3^ resolution. The extracted gray and white matter were nonlinearly registered to MNI space using SPM’s SHOOT algorithm. The gray matter was then warped using the individual flow fields resulting from the SHOOT registration, and voxel values were modulated for volumetric changed introduced by the high dimensional normalization to ensure that the total amount of gray matter volume was preserved. Images were then smoothed (FWHM: 4 mm) to account for anatomical variability across individuals and potential misregistration to facilitate statistical comparison. The NBM region was segmented using an atlas derived from post-mortem brains (23) using a probabilistic threshold of 50%, which is consistent with the suggested threshold for realistic NBM extraction (24). The individual volumes of gray matter, white matter, and cerebrospinal fluid were calculated by summing the voxel intensities in each tissue from the segmentation maps and multiplying by the voxel volumes. The total intracranial volume (TIV) was calculated as the sum of the gray matter, white matter, and cerebrospinal fluid volumes. The NBM volume was then normalized for each participant by dividing it by the TIV to account for head size differences.

##### Tractography Pipeline

Further preprocessing of the MRI data was completed using the FMRIB Software Library (FSL) (25).The tractography pipeline was designed based on methods previously shown to be successful for segmenting the two NBM tracts of interest in older adults with and without neurodegenerative disease (11, 14, 18).

The NBM was used as the seed region and the external capsule, and cingulum were included as waypoint regions for the lateral and medial tracts respectively. The brainstem, hemisphere, anterior commissure, and internal capsule were added as regions of avoidance. The same NBM region was used as in the volumetric analyses above. The internal capsule, external capsule, and cingulum were extracted from the Johns Hopkins University white matter atlas (26), the anterior commissure from FSL’s XTRACT atlas (27), and the brainstem from FSL’s First automated segmentation tool (28). The hemisphere mask was manually created in 2mm MNI space by identifying the midline of the brain in the coronal plane in FSLeyes. The transformation from diffusion to T1 space was calculated using a linear (FLIRT) 6 degree of freedom (dof) transformation. A 12 dof affine linear (FLIRT) transformation followed by a nonlinear (FNIRT) transformation was used to create the transformation from T1 to MNI space. The inverse of these transformations was computed and concatenated to define the transformation from MNI to diffusion space, which was then applied to all regions.

After estimating the diffusion parameters with bedpostX, probabilistic tractography was completed using the standard 5000 samples and 0.2 curvature threshold settings in probtrackX. A template for both the lateral and medial NBM tracts was created using the probabilistic tractography outputs from the healthy controls. All extracted tracts were converted to MNI space using the transformation from the previously mentioned region of interest registration. The tracts for each hemisphere were flipped and overlayed onto the contralateral hemisphere to create consistent templates across hemispheres. A template of both the lateral and medial tracts was created for each hemisphere that contained tracts present in at least a third of the healthy control participants. This threshold was selected based on visual inspection of the resulting tracts to ensure appropriate portions of the tracts remained in the template. The template tracts were then transformed back into the subject space and binarized to create participant specific tract masks, in both the healthy controls and disease groups (see Figure 1).

**Figure 1.**
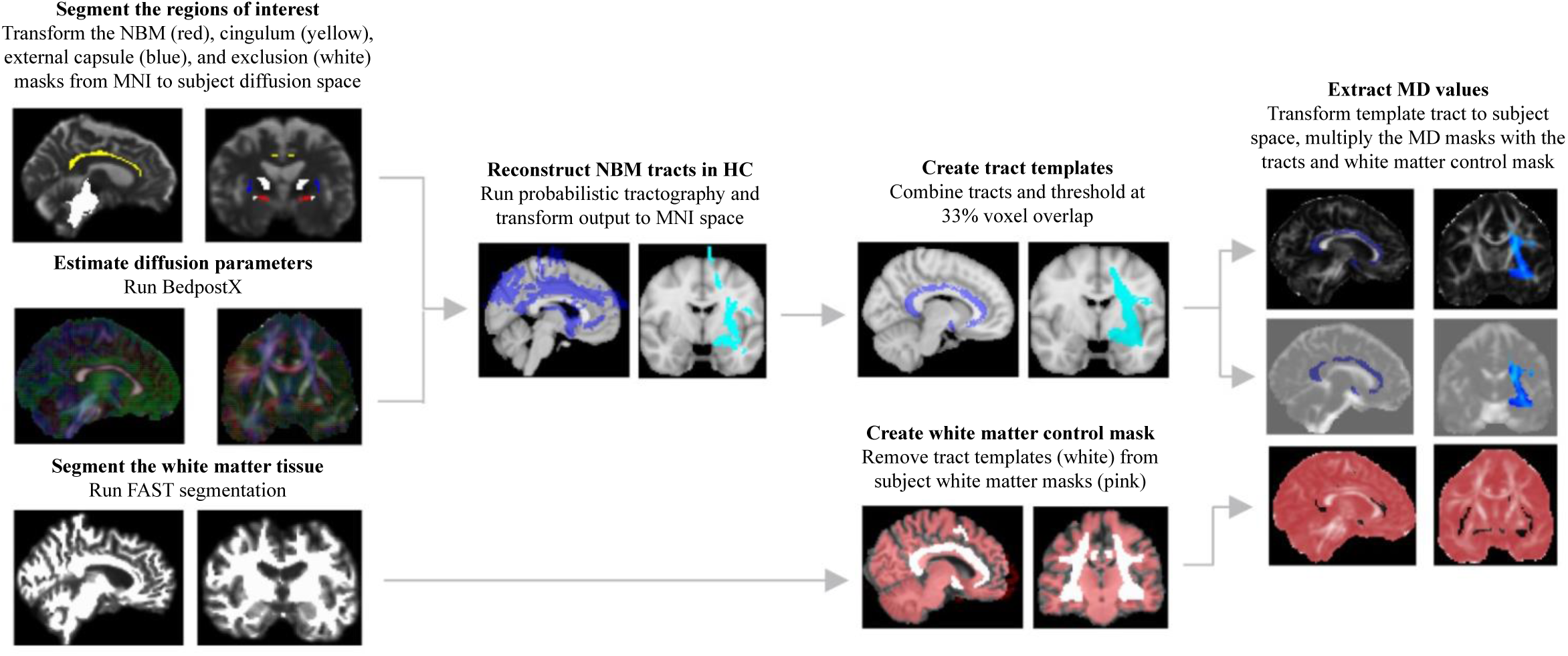
Flow of the tractography pipeline. Purple: medial tract; cyan: lateral tract; pink: white matter control mask.

The template tracts were then overlayed on to the ADNI preprocessed maps of mean diffusivity (MD) to determine the mean MD of each tract for every participant. A white matter control mask was created to determine if the differences in tract integrity may be reflective of brain-wide neurodegeneration rather than being tract specific. A white matter segmentation was first created using FSL FAST and the template tract was then registered and subtracted from this segmentation. This created a brain-wide map of the white matter excluding the tracts of interest. This control mask was then transformed back to diffusion space and the average MD values were extracted to be used as covariates in the analyses.

### 2.4. Statistical Analyses

All statistical analyses were completed in R version 4.2.2. Three analyses of linear mixed effects models were used to determine between group differences in NBM volume and MD of the lateral and medial tracts. NBM volume normalized by TIV was used for analyses with age and sex included as fixed effects and hemisphere specified as a random effect. For the MD analyses, the MD of the white matter control mask was also included as a fixed effect. Pairwise comparisons were completed using estimated marginal means, with the p-value adjusted using false discovery rate (FDR) to control for multiple comparisons (n=6 comparisons). Linear regression models were used to identify significant partial correlations between NBM volume and MD of the NBM tracts with each measure of cognitive function, controlling for age, sex, education, group, and MD of the white matter control mask for the tract integrity analyses. Significance was determined by an FDR adjusted alpha <0.05 (n=12 comparisons).

## 3. Results

The total sample (N=187) included 115 males (61%) with a mean (SD) age of 74.2 (7.0) years (see Table 1 for characteristics by group). There were no significant between-group differences in age or sex distribution. There were two people without cognitive data from the LMCI group and three in the AD group. Thus, there were a total of 182 people included in each of the linear regression models of cognition.

**Table 1.** Participant Characteristics.

|  | Mean (SD) or n (%) |  |  |  |
| --- | --- | --- | --- | --- |
|  | HC | EMCI | LMCI | AD |
| n | 50 | 50 | 37 | 50 |
| Age (yrs) | 74.1 (5.7) | 74.2 (7.3) | 72.8 (5.8) | 75.2 (8.5) |
| Sex (Male) | 27 (54) | 33 (66) | 25 (68) | 30 (60) |
| Education (yrs) | 16.6 (3.0) | 16.3 (2.6) | 16.3 (2.8) | 15.4 (2.9) |
| Executive Function | 0.67 (0.6) | 0.23 (0.7) | -0.03 (0.8) | -0.95 (0.9) |
| Memory | 1.09 (0.7) | 0.63 (0.8) | -0.34 (0.6) | -1.06 (0.6) |
| Language | 0.85 (0.7) | 0.47 (0.8) | 0.07 (0.7) | -0.81 (1.1) |
| Visuospatial | 0.19 (0.6) | 0.12 (0.6) | -0.20 (0.8) | -0.43 (0.8) |
| Total Intracranial Volume (mm<sup>3</sup>) | 1.39 (0.1) | 1.44 (0.2) | 1.43 (0.2) | 1.39 (0.2) |
| Control Mask MD | 0.00092 (0.00007) | 0.00092 (0.00006) | 0.00093 (0.00007) | 0.00097 (0.00007) |

### 3.1. Differences in NBM volume

There was a significant effect of group on NBM volume (F=49.05, p<0.0001, see Figure 2). Patients with AD had the smallest NBM volumes (p<0.0001 for all comparisons). LMCI also had smaller NBM volumes than the EMCI (p<0.0001) and HC (p<0.0001) groups. However, there was no significant difference between the EMCI and HC groups (see Table 2).

**Figure 2.**
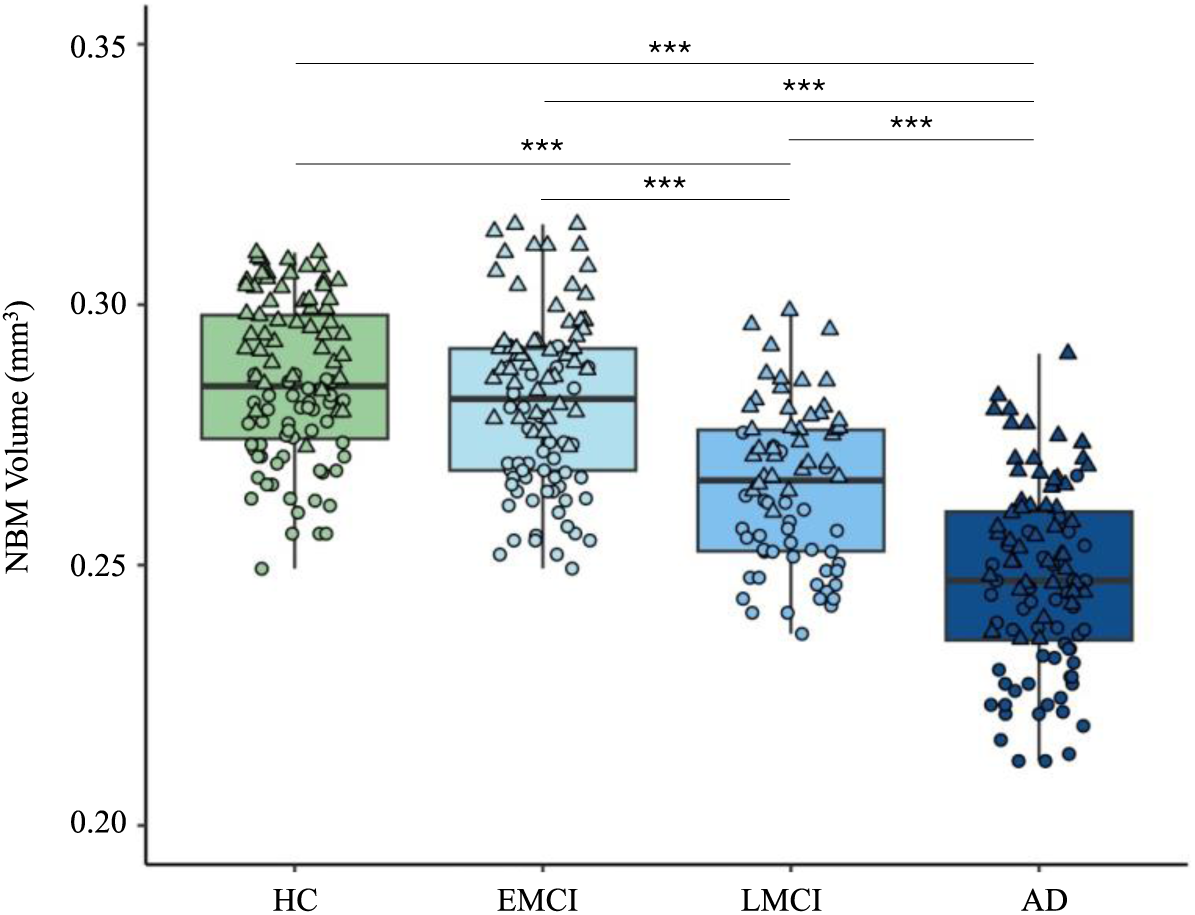
Comparison of NBM volume across healthy controls (HC), early mild cognitive impairment (EMCI), late mild cognitive impairment (LMCI), and Alzheimer’s disease (AD). Triangles: left hemisphere; Circles: right hemisphere. ***p<0.0001

**Table 2.** The estimated mean differences in tract MD and FDR-adjusted p-values of the pairwise comparisons.

|  | HC |  | EMCI |  | LMCI |  | AD |  |
| --- | --- | --- | --- | --- | --- | --- | --- | --- |
|  | Mean Difference<br>(95% CI) | *p* | Mean Difference<br>(95% CI) | *p* | Mean Difference<br>(95% CI) | *p* | Mean Difference<br>(95% CI) | *p* |
| **NBM Volume** |  |  |  |  |  |  |  |  |
| **HC** | - |  | 0.00265<br>(-0.00624, 0.0115) | 0.43 | 0.02057<br>(0.01090, 0.0302) | <0.0001*** | 0.03556<br>(0.02668, 0.0444) | <0.0001*** |
| **EMCI** | 0.00265<br>(-0.00624, 0.0115) | 0.43 | - |  | 0.01792<br>(0.00829, 0.0275) | <0.0001*** | 0.03290<br>(0.02403, 0.0418) | <0.0001*** |
| **LMCI** | 0.02057<br>(0.01090, 0.0302) | <0.0001*** | 0.01792<br>(0.00829, 0.0275) | <0.0001*** | - |  | 0.00365<br>(0.00540, 0.0247) | 0.0001** |
| **AD** | 0.03556<br>(0.02668, 0.0444) | <0.0001*** | 0.03290<br>(0.02403, 0.0418) | <0.0001*** | 0.00365<br>(0.00540, 0.0247) | 0.0001** | - |  |
| **Lateral Tract** |  |  |  |  |  |  |  |  |
| **HC** | - |  | -0.00002<br>(-0.00004, 0.0000007) | 0.02* | -0.00002<br>(-0.00004, 0.000003) | 0.03* | -0.00006<br>(-0.00008, -0.00004) | <0.0001*** |
| **EMCI** | -0.00002<br>(-0.00004, 0.0000007) | 0.02* | - |  | 0.0000004<br>(-0.00002, 0.00002) | 0.96 | -0.00004<br>(-0.00006, -0.00002) | <0.0001*** |
| **LMCI** | -0.00002<br>(-0.00004, 0.000003) | 0.03* | 0.0000004<br>(-0.00002, 0.00002) | 0.96 | - |  | -0.00004<br>(-0.00006, -0.00002) | <0.0001*** |
| **AD** | -0.00006<br>(-0.00008, -0.00004) | <0.0001*** | -0.00004<br>(-0.00006, -0.00002) | <0.0001*** | -0.00004<br>(-0.00006, -0.00002) | <0.0001*** | - |  |
| **Medial Tract** |  |  |  |  |  |  |  |  |
| **HC** | - |  | -0.00002<br>(-0.00005, 0.000009) | 0.09 | -0.00003<br>(-0.00006, -0.000002) | 0.008** | -0.00007<br>(-0.0001, -0.00004) | <0.0001*** |
| **EMCI** | -0.00002<br>(-0.00005, 0.000009) | 0.09 | - |  | -0.00001<br>(-0.00004, 0.00002) | 0.24 | -0.00005<br>(-0.00008, -0.00002) | <0.0001*** |
| **LMCI** | -0.00003<br>(-0.00006, -0.000002) | 0.008** | -0.00001<br>(-0.00004, 0.00002) | 0.24 | - |  | -0.00004<br>(-0.00007, -0.000006) | 0.004** |
| **AD** | -0.00007<br>(-0.0001, -0.00004) | <0.0001*** | -0.00005<br>(-0.00008, -0.00002) | <0.0001*** | -0.00004<br>(-0.00007, -0.000006) | 0.004** | - |  |

\*p<0.05; \*\*p<0.01; \*\*\*p<0.0001

### 3.2. Differences in integrity of the lateral and medial NBM tracts

There was a significant effect of group on the MD of both the lateral (F=23.14, p<0.0001) and medial (F=13.97, p<0.0001, see Figure 3) NBM tracts. Patients with AD had significantly greater MD in the lateral and medial tracts than the HC (p<0.0001), EMCI (p<0.0001) and LMCI (p<0.0001 for lateral and p=0.004 for medial tract) groups. LMCI showed significantly greater MD than HC in both the lateral (p=0.03) and medial (p=0.008) tracts. Notably, EMCI had significantly higher MD than HC in the lateral tract (p=0.02), but no significant difference was seen between these groups in the medial tract (see Table 2). There was no significant difference between the EMCI and LMCI for either tract. However, visual interpretation of the data suggests LMCI may be higher than EMCI in the medial tract (see Figure 3).

**Figure 3.**
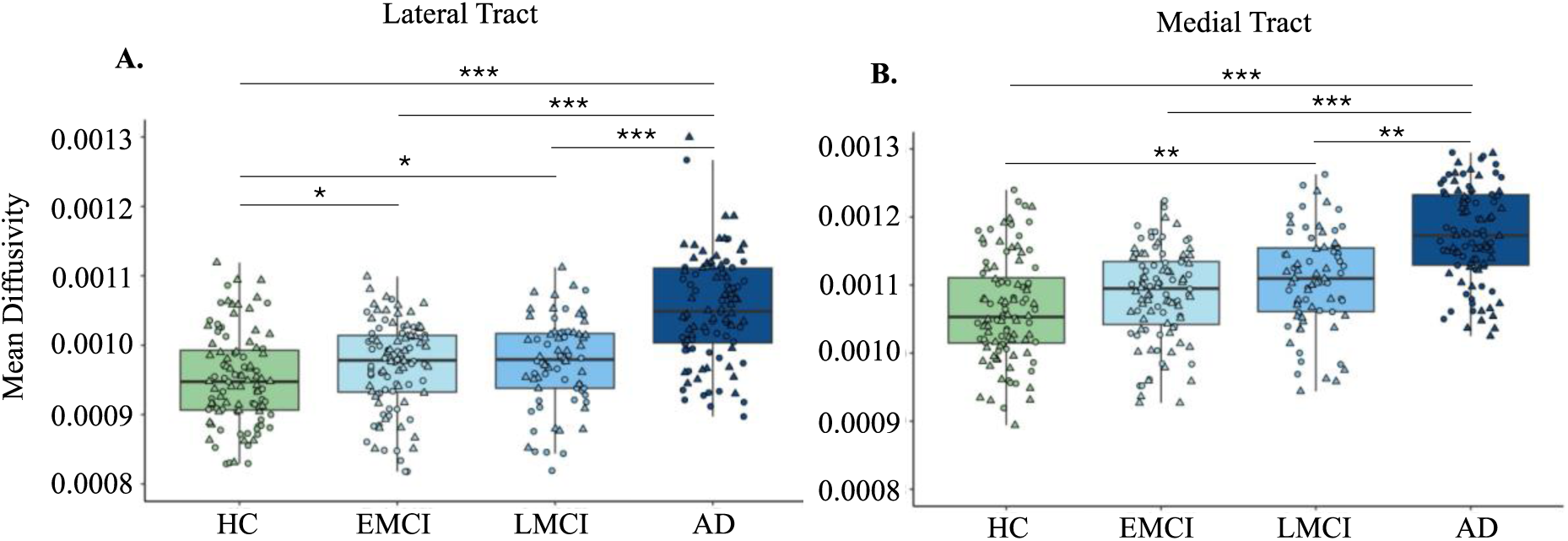
Comparison of the mean diffusivity of the lateral (A) and medial (B) NBM tracts across healthy controls (HC), early mild cognitive impairment (EMCI), late mild cognitive impairment (LMCI), and Alzheimer’s disease (AD). Triangles: left hemisphere; Circles: right hemisphere. *p<0.05; **p<0.01; ***p<0.0001

### 3.3. Associations with cognitive function

Greater MD of the lateral tract was significantly associated with poorer executive function (p<0.001) and language performance (p=0.022). In addition, while associations between NBM volume and MD of both the tracts with memory trended towards significance (unadjusted p<0.05), these associations did not survive after controlling for multiple comparisons. A trend was also seen between language and MD of the medial tract (unadjusted p<0.05). There was no association between NBM volume or the medial tract with executive function, or with any volumetric or tract integrity measures and visuospatial function (see Figure 4).

**Figure 4.**
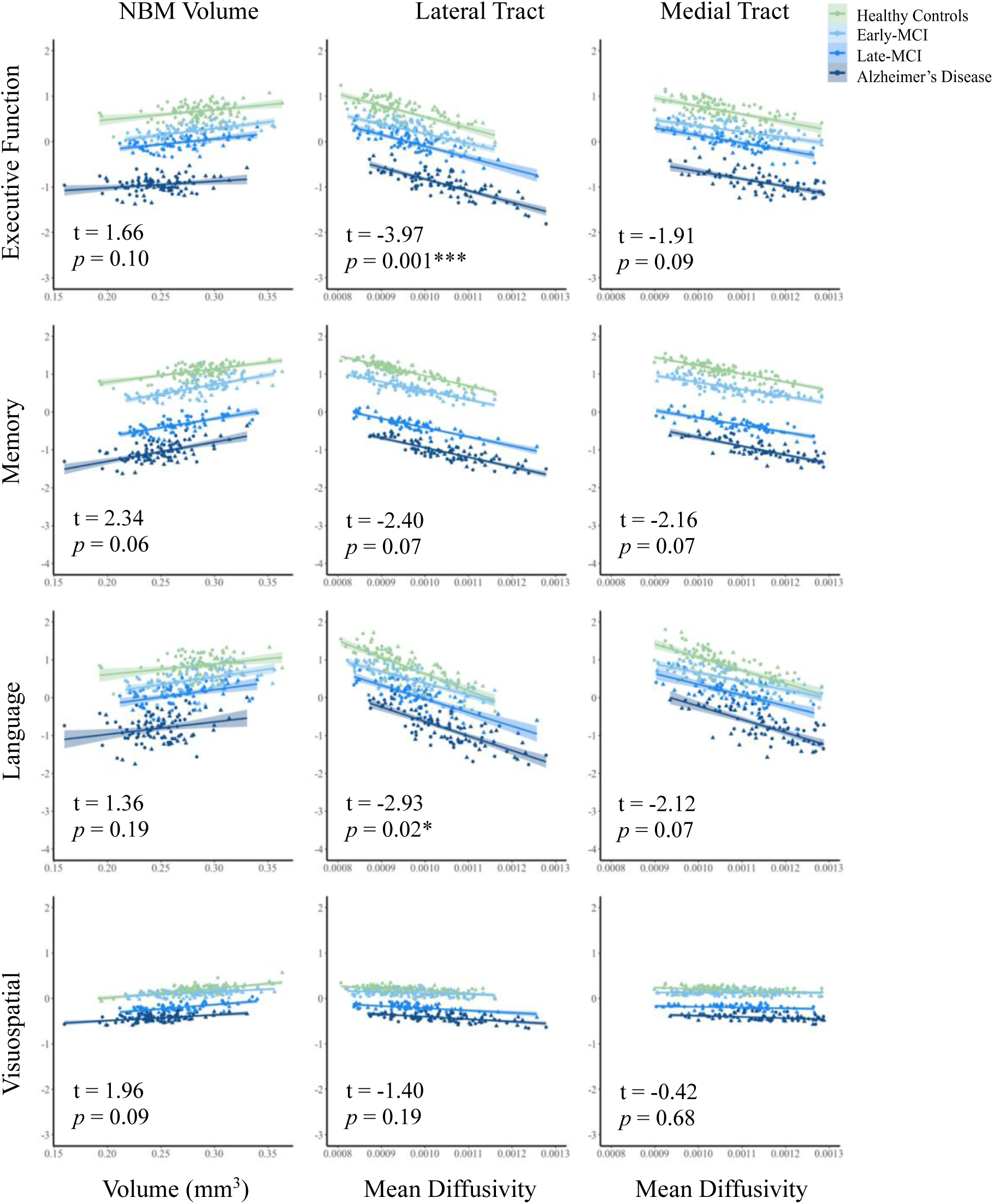
Predicted cognitive scores based on the linear regression models of normalized NBM volume, and mean diffusivity of both the lateral and medial NBM tracts. Triangles: left hemisphere; Circles: right hemisphere. p-values are FDR corrected. *p<0.5; **p<.001

## 4. Discussion

This study is the first to investigate differences in NBM volume and integrity of the NBM lateral and medial tracts in people with AD, LMCI, EMCI, and those who are cognitively healthy. We identified that NBM volume was significantly smaller in the AD group followed by those with LMCI compared to those with EMCI and HC. The AD group also had significantly lower integrity of both tracts compared to the MCI and HC groups. We found no difference in integrity between the EMCI and LMCI groups in either tract. However, while both MCI groups had significantly lower integrity of the lateral tract compared to the HC group, only the LMCI group had significantly lower integrity of the medial tract than the HC. No difference was identified between the HC and EMCI groups for this tract. Finally, we observed significant associations between the integrity of the lateral tract with performance in executive function and language tasks. No association was identified between any of the cognitive domains and NBM volume or the integrity of the medial tract.

It is notable that while the AD and LMCI groups had significantly smaller NBM volumes, there was no significant difference between the EMCI and HC groups. Differences were observed here in the integrity of the lateral tract, suggesting that it may be more sensitive to disease stage than NBM volume as a whole. This is consistent with previous research identifying greater ability of NBM tract integrity to discriminate between disease stages than volumetric measures at earlier stages of disease (14, 29). Previous research from the ADNI database has found differences in NBM volume between EMCI and HC groups when using upsampling for greater MRI resolution but not when using the original participant images (10). Thus, the ability to detect more subtle differences between earlier stage MCI and HC in volumetric NBM measures may require greater scanning resolution than lateral tract integrity, which is likely more robust.

The trajectory of the lateral tract integrity with disease stage is consistent with previous findings that used a separate cohort (14). However, differences in the integrity of the medial tract were previously only identified between the AD and HC groups. Our findings differ in the identification of an additional difference between the LMCI and HC groups in the medial tract. This may be because the previous studies used an all-encompassing MCI group as opposed to differentiating between those at early and late stages of decline. This demonstrates the need for more sensitive approaches to detect pathological differences across the earliest stages of disease. Taken together, our results provide substantial evidence that reduced integrity of the NBM tracts should be considered a potential biomarker for cognitive decline in neurodegenerative disease, with a particular relevance of the lateral NBM tract at earlier disease stages.

Previous findings identified the involvement of the lateral tract in executive function, memory, and language in older adults (11) and those with vascular cognitive impairment (12). Our findings support this and are the first to extend these associations to people with Alzheimer’s-related MCI and AD. While it was previously shown that integrity of the lateral tract was also associated with attention in this population (14), we were not able to assess this directly. However, it is widely accepted that attention is required for cognitive tasks involving executive function (30). Thus, it is possible to imply that our findings may support this relationship as well.

Trends were identified between NBM volume and integrity of the medial tract with memory and language (medial tract only). However, these associations did not survive after adjusting for multiple comparisons. A lack of association between these metrics and cognitive function is consistent with previous evidence in patients with MCI, AD, dementia with Lewy bodies, and Parkinson’s disease (11, 14, 18). Our findings support the notion that the lateral NBM tract specifically, has importance for cognitive function, while the role of the medial tract remains ambiguous. The NBM cholinergic system also plays a role in sleep, emotion, and psychiatric symptoms (31). Greater investigation is needed to determine whether the medial tract may therefore affect cognitive function through an indirect involvement in these systems.

The lack of association between NBM volume or tract integrity and visuospatial function is difficult to interpret due to limitations in the tasks used, which required the drawing of pictured items. While categorised as visuospatial function, these tasks are heavily confounded by manual reconstruction ability and have a limited scoring range (0-1 for each item), compared to other metrics such as the Benton judgment of line orientation that requires only verbal responses (32). However, previous research using the potentially preferred metrics still only identified associations between NBM volume (33), but not tract integrity (18), with visuospatial function in patients with Parkinson’s disease. Therefore, it is also possible that there is limited relevance of the NBM tracts to this cognitive domain. Further research with a range of appropriate metrics is needed to assess the role of the NBM and NBM tract integrity in visuospatial function.

This study is not without limitations. Due to study designs beyond our control, we were limited in the categorisation and options of cognitive domains available for evaluation. While we were able to include a wide range of the categories of interest, it is possible that some tasks, such as those in the visuospatial category, limited the ability to perform effective analyses. The NBM is very small region at approximately 13-14mm in the anterior-posterior direction and 16-18mm in the medio-lateral direction (34). As NBM volume was segmented in 2mm^3^ resolution, the accuracy of the extracted volumes may be suboptimal. Further, this study was cross-sectional meaning we were not able to evaluate longitudinal changes in people who may change diagnosis categories over time. Thus, although we can infer the potential disease trajectory based on our comparison across groups, further investigation comparing within patient changes would provide considerable support to our findings.

### 4.1. Conclusion

Refining MCI into early and late-stage disease is beneficial for identifying differences in white matter integrity compared to healthy controls. In addition, reduced integrity of the lateral NBM tract is most sensitive to the earliest signs of MCI and is associated with reduced executive function and language performance. Thus, the lateral NBM tract should be considered an important therapeutic target for early detection and intervention in AD.

## Data Availability

Data used in preparation of this article were obtained from the Alzheimer's Disease Neuroimaging Initiative (ADNI) database (adni.loni.usc.edu).

https://adni.loni.usc.edu

